# Paclitaxel-Coated or Uncoated Devices: Significant Differences in Patient Populations and Mortality Led to Study Incomparability

**DOI:** 10.1101/2021.01.29.21250732

**Authors:** Chenyang Zhang, Guosheng Yin

## Abstract

The SWEDEPAD trial reported an unplanned interim analysis to show no difference in the mortality rate between the paclitaxel-coated and uncoated groups (Nordanstig et al., 2020), which contradicts the long-term risk of paclitaxel-coated devices claimed by a meta-analysis (Katsanos et al., 2018). However, there existed significant differences in mortality rates between the SWEDEPAD trial and the trials included in the meta-analysis, which were caused by significant differences in the patient populations. As a result, the SWEDEPAD trial and meta-analysis results are not directly comparable. An updated meta-analysis including the SWEDPEPAD trial and all studies in the meta-analysis (Katsanos et al., 2018) shows marginal differences in mortality rates between the paclitaxel-coated and control groups at two years with Bayesian relative risk (RR) 1.39 (95% credible interval (CrI) [1.01, 2.39]) and frequentist RR 1.16 (95% confidence interval (CI) [0.99, 1.36]) and differences in mortality rates during the entire follow-up period with Bayesian RR 1.29 (95% CrI [1.01, 1.72]) and frequentist RR 1.13 (95% CI [0.99, 1.28]) under random-effects models. Given the relatively short follow-up thus far in the SWEDEPAD trial (with a mean follow-up of 2.49 years) and the paclitaxel-coated risk being long-term (e.g., 4 or 5 years), the interim results on the risk of paclitaxel-coated devices reported by the SWEDEPAD trial warrant further investigation.

## 1 Introduction

The SWEDEPAD trial^1^, which are investigating paclitaxel-coated versus uncoated devices in patients with peripheral artery disease (PAD), was temporarily suspended on December 2018 due to the potential long-term risk of paclitaxel-coated devices reported by a meta-analysis.^2^ An unplanned interim analysis of the SWEDEPAD trial showed no difference in the mortality rate between the paclitaxel-coated and uncoated groups at one year with a hazard ratio (HR) of 1.03 (95% CI [0.77, 1.37]) or during the entire follow-up period with an HR of 1.06 (95% CI [0.92, 1.22]). Based on these results, it was decided to resume enrollment in the SWEDEPAD trial on March 23, 2020. However, Figure 2 of the SWEDEPAD trial publication shows that the cumulative incidence curves of drug-coated devices constantly stayed above those of uncoated either for the overall population or subgroups of chronic limb threatening ischemia or intermittent claudication.^1^ The risk of paclitaxel-coated devices is known to be of long term,^2,3^ while the SWEDEPAD trial has so far had a mean follow-up of 2.49 years which is still relatively short for evaluating long-term risk.

## 2 Methods and Results

### 2.1 Baseline Differences in Patient Populations

By pooling the numbers of deaths and patients from all studies included in the meta-analysis, we treat the combined data as a mega-trial,^2^ and compare the differences in mortality between the SWEDEPAD trial^1^ and mega-trial. For the paclitaxel-coated group, we found significant differences in the all-cause death rates at one year with relative risk (RR) 4.4 (95% CI [3.2, 6.0]) and during the entire study period with RR 3.7 (95% CI [3.1, 4.5]). For the uncoated group (control), the mortality rate differences between the SWEDEPAD trial^1^ and mega-trial were also significant with RR 4.2 (95% CI [3.0, 5.9]) at one year, and RR 5.7 (95% CI [4.5, 7.2]) during the entire study period. All RRs comparing the SWEDEPAD trial^1^ and mega-trial^2^ are larger than 3.5 with p-values smaller than 0.0001, which could have attributed to incomparability between the SWEDEPAD trial^1^ and the trials included in the meta-analysis^2^. There are significant differences at the baseline in the patient populations between the SWEDEPAD trial and trials in meta-analysis: the former had 35% intermittent claudication (IC) and 65% chronic limb threatening ischemia (CLTI) while the latter had 89% IC and 11% CLTI. CLTI is an advanced stage of PAD. Compared with intermittent claudication, CLTI has a negative prognosis within a year after the initial diagnosis, with a 1-year amputation rate of approximately 12% and mortality rates of 50% at 5 years and 70% at 10 years.^4^ The baseline prognosis difference could be a main cause of observed differences in mortality rates between the studies.

### 2.2 Updated Meta-Analysis

We conducted an updated meta-analysis which included the SWEDEPAD trial^1^ and all studies in the meta-analysis^2^. The number of deaths by two years of the SWEDEPAD trial^1^ was estimated by multiplying the estimated two-year cumulative incidence rate and the total number of patients. For each study in the meta-analysis^2^, we took the results with the longest follow-up as those for the entire study period. The R packages ‘meta’ and ‘bayesmeta’ were used to formulate frequentist and Bayesian random-effects models, respectively.

As shown in Table 1, both frequentist and Bayesian random-effects models demonstrate no difference in one-year all-cause death rates between paclitaxel-coated and uncoated arms, indicating no short-term risk of paclitaxel-coated devices. At two years or during the entire follow-up period, the frequentist random-effects model yields marginally insignificant differences in all-cause death rates with two-year RR 1.16 (95% CI [0.99, 1.36], P=0.063), and the entire follow-up RR 1.13 (95% CI [0.99, 1.28], P=0.065). However, the Bayesian random-effects model yields an RR of 1.39 (95% credible interval (CrI) [1.01, 2.39]) at two years and an RR of 1.29 (95% CrI [1.01, 1.72]) during the entire follow-up period, which suggests significant risk of paclitaxel-coated devices with longer-term follow-ups. The posterior probabilities for the death rate of the paclitaxel-coated arm being higher than that of the control arm were around 0.98 at two years and during the entire follow-ups, with Bayes factors larger than 40. This provides strong evidence for the long-term mortality risk of paclitaxel-coated devices.

**Table 1.** Relative risk and interval estimates of the meta-analysis including the SWEDEPAD trial^1^ and all studies in the meta-analysis^2^.

|  |  | Follow-up period |  |  |
| --- | --- | --- | --- | --- |
|  |  | One year | Two years | Entire follow-up |
| Frequentist | Random-effects | 1.04 [0.84, 1.28] * | 1.16 [0.99, 1.36] | 1.13 [0.99, 1.28] |
|  | P value | 0.713 | 0.063 | 0.065 |
| Bayesian | Random-effects | 1.04 [0.77, 1.41] | 1.39 [1.01, 2.39] | 1.29 [1.01, 1.72] |
|  | Posterior probability<br>of (RR>1) | 0.613 | 0.978 | 0.980 |

\*Relative risk (RR) with 95% confidence interval (frequentist) or equal-tailed credible interval (Bayesian) in the brackets.

## 3. Conclusion

The SWEDEPAD trial^1^ enrolled 2289 patients, which is about half of the total number of patients included in the meta-analysis^2^. Although one study with such a large sample size and insignificant results was added, the new meta-analysis still yields marginally significant differences in the mortality rates at two years and during the entire follow-up. Moreover, with a mean follow-up of only 2.49 years, the results of the SWEDEPAD trial warrant further investigation, because the risk from paclitaxel-coated devices is long-term.^5^

## Data Availability

The data that support the findings of this study are available from the corresponding author, G Yin, upon reasonable request.

